# Differentiating nonfluent/agrammatic and logopenic primary progressive aphasia in Catalan-Spanish bilinguals by applying multilingual multimodal machine learning (M³L) to connected speech

**DOI:** 10.64898/2026.09.18.26363435

**Authors:** Lokesha S. Pugalenthi, Andrew P. Collins, Núria Montagut Colomer, Sonia-Karin Marqués-Kiderle, Camille A. Wagner Rodriguez, Jan Christian Holst Chaires, Whendy Avila Motta, Júlia Filella-Mercè, Junyi Jessy Li, Fernando Llanos, Nuole Zhu, Sara Rubio-Guerra, Ignacio Illan-Gala, Sergi Borrego-Écija, Albert Lladó, Juan Fortea, Alberto Lleó, Raquel Sánchez-Valle, Maya L. Henry, Miguel Ángel Santos Santos, Stephanie M. Grasso

**Affiliations:** Department of Electrical and Computer Engineering, Rice University, Houston, Texas, USA; Department of Speech, Language, and Hearing Sciences, The University of Texas at Austin, Austin, Texas, USA; Department of Speech, Language, and Hearing Sciences, University of Connecticut, Storrs, Connecticut, USA; Alzheimer’s disease and other Cognitive Disorders Unit, Service of Neurology, Hospital Clínic de Barcelona, Barcelona, Spain; Sant Pau Memory Unit, IR SANT PAU, Hospital de la Santa Creu i Sant Pau, Barcelona, Spain; Department of Linguistics, The University of Texas at Austin, Austin, Texas, USA; Centro de Investigación Biomédica en Red en Enfermedades Neurodegenerativas (CIBERNED), Madrid, Spain; Department of Neurology, Dell Medical School, The University of Texas at Austin

**Author notes:** Indicates that Stephanie M. Grasso and Miguel Angel Santos Santos share senior authorship.

**Keywords:** differential diagnosis, subtyping, natural language processing, ensemble modelling, discourse

## Abstract

**Background:** The nonfluent/agrammatic (nfv) and logopenic (lv) variants of primary progressive aphasia (PPA) disrupt fluency through distinct underlying neurocognitive mechanisms. Differential diagnosis currently requires hours of cognitive-linguistic testing, with additional barriers for bilingual patients due to a shortage of bilingual service providers and a lack of well-established assessment methods. In English speakers, a promising automated approach for differentiating nfvPPA and lvPPA is to derive speech-timing measures and linguistic features from connected speech as input to machine learning (ML) classification algorithms. To our knowledge, this approach has not been evaluated in the context of bilingualism.

**Methods:** Thirty-four Catalan-Spanish simultaneous bilingual patients (lv = 24, nfv = 10) were asked to describe a picture (Western Aphasia Battery Picnic Scene) in both their dominant and non-dominant language. From the participant’s recorded response, we derived four feature sets: speech-timing measures, derived with PRAAT; word-level parameters, derived from corpora; linguistic features, derived with the natural language processing tools SpaCy and CLAN; image-text congruence scores, derived with the vision-language encoder Multilingual-CLIP. Each feature set was fed into classification algorithms for differentiating nfv from lv in participants’ non-dominant and dominant samples. Then, we combined each feature set’s classifier into an ensemble model. We used the McNemar test to determine the statistical significance of differences in classification performance between responses in the non-dominant and dominant language.

**Results:** The best-performing classifier achieved F1 macro scores of 93% (word-level parameters) and 92% (ensemble) in the non-dominant and dominant language, respectively. For all feature sets and ensemble models, classification performance did not significantly differ between the non-dominant and dominant language. Ensemble modeling did not significantly improve classification performance in either language.

**Conclusions:** Taking advantage of recent advances in multilingual multimodal machine learning, we accurately differentiate Spanish-Catalan bilingual individuals with nfvPPA and lvPPA using a largely automated, time-efficient (1–2 minutes), and ecologically valid connected-speech-based approach. Future directions include evaluating this approach on larger datasets balanced by PPA subtype, using automated transcriptions of connected speech. Our study represents a step towards addressing current inequities in PPA differential diagnosis for non-English-speaking bilingual speakers.

**Trial registration:** Data from the clinical trial NCT05741853 was retrospectively analyzed

## Background

Primary progressive aphasia (PPA) is a neurodegenerative syndrome characterized by the gradual erosion of language and/or speech^1^. Although symptoms initially concentrate in speech and language, cognitive symptoms emerge with disease progression^1^. PPA is categorized into three variants, each with a unique pattern of speech and/or language deficits. The semantic variant (svPPA) is associated with a loss of core semantic knowledge, leading to deficits in word retrieval and comprehension, atrophy observed in the anterior temporal lobes (left > right)^1^, and is most often associated with transactive response DNA-binding protein 43 type C pathology (TDP-43-C)^2^. The logopenic variant (lvPPA) is associated with impaired phonological processing, leading to deficits in word retrieval and repetition, atrophy observed in the left temporoparietal cortex^1^, and is most often associated with underlying Alzheimer’s disease pathology^2^. Lastly, the nonfluent/agrammatic variant (nfvPPA) is associated with impaired grammar and/or motor speech impairment, atrophy of the left fronto-insular region^1,3^, and is most often associated with underlying tau pathology^2^.

Fluency is a multidimensional construct, reflecting contributions from motor speech, syntax, word finding, phonology, and prosody. Compared to individuals with svPPA, individuals with nfvPPA and lvPPA produce less fluent speech for different underlying reasons. Thus, distinguishing between nfvPPA and lvPPA is particularly challenging^4^. For example, apraxic speech-sound errors, typically seen in nfvPPA, can be difficult to distinguish from phonological paraphasias^5^. Although the source of impaired fluency in nfvPPA and lvPPA differs (motor speech and/or grammar vs. phonological processing and/or word retrieval, respectively), these two PPA subtypes can look similar, particularly in the mild, early stages of disease progression^6^.

The inherent characteristics of *bilingual* speech and language further complicate PPA subtyping. For example, bilinguals perform less favorably compared to monolinguals on tasks associated with word retrieval^7^. In addition, bilinguals have been shown to have greater tip-of-the-tongue states relative to monolingual speakers^7^. Therefore, diagnosing PPA and its variants in bilingual speakers is particularly challenging, as select PPA symptoms overlap with typical features of speech production in bilinguals. Beyond these speech-language characteristics of bilinguals, the accessibility of evidence-based services also proves challenging. More specifically, although most of the world’s population is bilingual, a shortage of bilingual service providers may be found^8^. Irrespective of sociocultural context, methods for PPA assessment in bilinguals have yet to be thoroughly investigated^9,10^.

For monolinguals and bilinguals alike, differential diagnosis by PPA subtype relies on comprehensive cognitive-linguistic assessment administered and interpreted by experienced clinicians in routine clinical care^11^. These assessments typically require overt responses and often comprise hours of tasks (e.g., naming pictures of objects), often leading to fatigue, which may negatively affect the validity and reliability of the testing results^11^. These somewhat artificial tasks may not accurately assess one’s ability to partake in everyday conversations^12^. Given these challenges, clinicians and researchers seek alternative or complementary tools for confirming a diagnosis^13^.

To address these challenges in differentiating nfvPPA and lvPPA in bilingual individuals specifically, we developed a largely automated tool by deriving *speech-language features* from elicited connected speech as input to multivariate classification algorithms. This methodology requires only 1-2 minutes of connected speech production, which is significantly less time-consuming than traditional assessments. Additionally, this method is less expensive and/or invasive than deriving PPA-implicated biomarkers from blood, cerebrospinal fluid, and neuroimaging^13–15^ and has greater ecological validity than traditional assessment methods.

In monolingual English speakers with PPA, multiple studies have used analysis of connected speech for differential diagnosis^16–20^. Fraser et al.^17^ was the first study to evaluate this methodology in PPA, achieving 79% accuracy in differentiating nfvPPA and svPPA (N=24) solely using linguistic features. Rezaii et al.^19^ differentiated four groups (controls, svPPA, lvPPA, nfvPPA; 98% accuracy, N=98) using linguistic features. Speech-language features have also been used together to differentiate all three PPA subtypes in monolingual English speakers (80% accuracy, N=44)^20^. Of most relevance to the current study, Lukic et al. is the only study, to our knowledge, to have used this methodology to differentiate solely between nfvPPA and lvPPA (82% area under the curve, N=61)^18^ in monolingual English speakers.

Outside of monolingual English speakers, two studies have used this approach for differential diagnosis of PPA subtype. In monolingual Spanish speakers with PPA, speech-language features have been used together to differentiate all three PPA subtypes (78% accuracy, N=87)^21^. In monolingual Italian speakers with PPA, speech-language features have been used together to differentiate nfvPPA from lvPPA (82% accuracy, N=61), as well as differentiate all three PPA subtypes (64% accuracy, N=84)^22^. However, no study has evaluated this approach in bilingual individuals with PPA, let alone evaluated whether language dominance affects classification performance.

In the current study, focused on bilingual speakers with PPA, we derived four feature sets using natural language processing (NLP): (1) *speech-timing measures*, (2) *linguistic features*, (3) corpora-derived *word-level parameters*, and (4) *image-text congruence scores*. The last is the most novel feature set, measuring semantic congruence between a transcription and a picture using vision–language encoders^23^. Unlike previous automated PPA subtyping work, which has focused solely on microlinguistic features, our image-text congruence classifier captures macrolinguistic discourse-level processes. For each feature set, we built classifiers using each patient’s connected speech sample in (a) their non-dominant language (NDL), (b) dominant language (DL).

First, we evaluated classification performance of each feature set in both the NDL and DL. Our first hypothesis is that all feature sets, regardless of language, would outperform a naïve classifier that predicts every sample as the majority class in our dataset (lvPPA). Our second hypothesis is that classification performance would be greater in the NDL because previous work indicates that the NDL is more susceptible to initial PPA symptoms (i.e., greater observable deficits in the NDL)^24^. If this is true, this would imply clinicians should prioritize testing in the NDL, despite clinician’s tendency to test in the DL^10^. Second, we evaluated whether ensemble modeling provides a more holistic and accurate assessment of connected speech than individual feature-set classifiers. We implemented a late-fusion^25^ ensemble model that combined lvPPA prediction probabilities from each feature set’s classifier. We hypothesized that the ensemble would outperform each feature set’s classifier, implying that simultaneously considering multiple aspects of language is critical for accurate classification. Third, we identified the most discriminative features in the NDL and DL, and compared how each feature set differs in their contribution to ensemble classification performance. In sum, the purpose of the current study was to differentiate between the nonfluent/agrammatic (nfvPPA) and logopenic (lvPPA) variants in bilinguals (Catalan-Spanish) with PPA by deriving speech-language features from connected speech as input to classification algorithms.

## Methods

### Participants

Thirty-four Catalan-Spanish simultaneous bilingual individuals with PPA (lvPPA = 24, nfvPPA = 10; see Table 1) participated in the current study. Participants were recruited through an ongoing clinical and research collaboration. The study was approved by the ethics committees of Hospital de Sant Pau and Hospital Clínic de Barcelona, as well as the Institutional Review Board at the University of Texas at Austin. All participants provided written informed consent. Individuals included in this study participated in a larger study (NCT05741853), which required participants to have a baseline MMSE of 10 or greater, and not to have any uncorrected hearing or visual impairments. Participants were diagnosed by behavioral neurologists specialized in atypical dementias, and all participants met consensus criteria for clinical diagnosis of either lvPPA or nfvPPA^1^. Most participants underwent cerebrospinal fluid (CSF) biomarker testing. For the 22 of 24 individuals with lvPPA that underwent CSF biomarker testing, all were confirmed to be amyloid-positive. For the eight out of 10 individuals with nfvPPA that underwent CSF biomarker testing, all were confirmed to be amyloid-negative. Amyloid status was determined in accordance with established cut-off values (Ab1−42 / pTau181 < 15.1)^26^. Individuals included in this study participated in a larger study, which required participants to have a baseline MMSE of 10 or greater, and not to have any uncorrected hearing or visual impairments.

**Table 1.** Participants’ Demographic and Cognitive-Linguistic Characteristics (Mean and (Standard Deviation)). Assessment scores all represent performance in the dominant language.

| Measure/Variable | lvPPA (N = 24) | nfvPPA (N = 10) | p-value |
| --- | --- | --- | --- |
| Age | 74 (6.61) | 77 (6.74) | .21 |
| Education | 14.5 (4.3) | 15 (4.6) | .77 |
| Sex (%; M= male) | 54.2% M | 70% M | .64 |
| Catalan Dominant<sup>27</sup> (%) | 75% | 50% | .31 |
| Mini-Mental State Exam<sup>28</sup> | 23.71 (5.07) | 27.33 (1.58) | .004 |
| QAB<sup>29</sup>/WAB<sup>30</sup> (/10) | 8.03 (1.04) | 8.15 (1.18) | .80 |
| MINT<sup>31</sup> (/32) | 19.13 (6.96) | 26.40 (5.68) | .005 |
| Apraxia Severity Rating<sup>32</sup> (range 0-7) | .07 (.27) | 2.38 (0.52) | <.0001 |
| Dysarthria Severity Rating<sup>32</sup> (range 0-7) | .07 (.27) | 1.25 (1.28) | .04 |
| Sentence Production Task (simple and complex sentences)<sup>33</sup> | 7.73 (4.43) | 9.75 (4.74) | 0.33 |
| Amyloid Positivity<sup>26</sup> (%) | 100% | 0% | N/A |

<sup>27</sup> Birdsong et al. (2012)
<sup>28</sup> Folstein et al. (1975)
<sup>29</sup> Wilson et al. (2018)
<sup>30</sup> Kertesz (2007)
<sup>31</sup> Gollan et al. (2012)
<sup>32</sup> Wertz et al. (1984)
<sup>33</sup> Varkanitsa et al. (2024)
<sup>26</sup> Alcolea et al. (2019)

### Language Dominance

Language dominance was determined using a comprehensive, previously validated self-report measure, the Bilingual Language Profile^27^. This tool assesses language dominance by assigning weights to responses across several measures: language history, language use, language proficiency, and language attitudes. From these weighted scores, a dominance index is derived, enabling identification of language dominance, as reported in Table 1.

### Connected Speech Task Materials and Administration

Each participant was asked to describe the picnic scene from the Western Aphasia Battery-Revised (WAB-R)^30^ in both their dominant and non-dominant language, with the following prompt administered in either language: “Tell me everything you see in the picture. Try to tell me in complete sentences.” The participant spoke uninterrupted during this task.

### Preprocessing

Each participant’s picture description was recorded in both their dominant and non-dominant language, either in person or via Zoom. All samples were reviewed for audio quality to ensure adequate sample quality (see Supplemental Materials, Section: Validation of linguistic and speech-timing feature extraction). Subsequently, rVAD^34^ was used to remove silence at the start and end of audio samples. Then, a transcript was generated for each preprocessed recording using the Whisper large-v3 model^35^. The transcripts were then manually corrected, segmented by utterance, and reformatted into a chat file using CLAN conventions by trained speech-language pathologists^36^. For a subset of samples (at least 20% in each language), inter-rater reliability was conducted using the RELY function in CLAN. In both Spanish (92% utterances; 89% words) and Catalan (92% utterances and 86% words), inter-rater reliability was high.

### Features derived

From our samples, we derived four feature sets: *linguistic features* (N=11), corpus-derived *word-level parameters* (N=9), *speech-timing measures* (N=9), and *image-text congruence scores* (N=5). Each feature’s definition and source are stated in Supplementary Table 1. We derived the following linguistic features^37,38^: number of words, proportion of fillers, type-token ratio, noun-to-verb ratio, propositional density, open-to-closed words ratio, average sentence length, subordination index, retracings, whole-word repetitions, and part-word repetitions. The following linguistic features were included because they were reported to be discriminative in previous studies of connected speech in PPA^17,21,39^: fillers^17^, type-token ratio^17^, noun-to-verb ratio^17^, average sentence length^21^, propositional density^39^, open-to-closed words ratio^39^. Subordination index and retracing were also included because reduced syntactic complexity is typically observed in nfvPPA^3^, and because individuals with lvPPA use increased circumlocutions to compensate for word-retrieval difficulties^40^. Part-and whole-word repetitions were also included to capture disruptions to fluency present in nfvPPA and lvPPA. The only features that are manually hand-coded by transcribers were retracings, part-and whole-word repetitions, all of which were derived with CLAN^36^.

We derived the following word-level parameters from extant corpora^41–46^: word frequency per million, verb frequency per million, noun frequency per million, percent of high frequency verbs, word length by syllables, word length by phonemes, concreteness, and age of acquisition. All but two word-level parameters were included because they were reported to be discriminative in previous studies of connected speech in PPA^16,17,19,21^. Word length by syllables and word length by phonemes were also included due to the phonological deficits present in lvPPA. As word length by syllables is less variable than word length by phonemes, including both parameters allows for the evaluation of two distinct levels of production and to account for the possibility of speakers producing words with the same number of syllables but different number of phonemes e.g. “casar” and “cantar.”

We derived the following speech-timing measures^47,48^: speech rate (number of syllables per second), number of words/minute, average syllable duration, articulation rate (number of syllables divided by phonation time, reflecting motor speech function more purely than speech rate), speech-pause ratio, number of pauses, mean pause duration, variance of pause duration, and number of propositions/minutes. All speech-timing measures were included because they were reported to be discriminative in previous studies of connected speech in PPA^20,21,39,48–52^.

We derived image-text congruence, a measure of semantic congruence between a text and a picture^23^, using multilingual vision-language encoders^54^, because we predicted that individuals with lvPPA would have lower scores due to anomia. In previous work with similar clinical groups, image-text congruence differentiated nfvPPA from svPPA^55^. We derived image-text congruence scores from multiple vision-language encoders, based on the intuition that combining their outputs would yield better results^56^.

To see each feature’s definition, source, and validation, please see Supplementary Tables 1-6 describing our procedures for validation of linguistic and speech-timing feature extraction.

### Classification algorithms

For each feature set, we fed its features into classification algorithms to differentiate nfvPPA from lvPPA separately for NDL and DL. We used classification algorithms from the ScikitLearn Python package.

We follow previous connected speech studies in reporting outcomes across multiple classification algorithms^17,20,21^ due to the lack of consensus on the optimal classification algorithm given speech-language features. We used classification algorithms that demonstrated high accuracy for this classification task in monolingual English and Spanish speakers, given *speech-timing measures* and *linguistic features* derived from connected speech: support vector machine^17,20,21^, feedforward neural network^20^, Decision Tree^20,21^, gradient boosting^18^. Before evaluating the classification algorithms of the support vector machine and feedforward neural network, we z-scored our data because these distance-based classification algorithms assume that each feature is on the same scale. For the support vector machine classification algorithm, principal component analysis was applied after z-scoring, with the number of principal components treated as a hyperparameter to be tuned. For the feedforward neural network classification algorithm, we used only one hidden layer to avoid overfitting, given the small size of our dataset. Since our feedforward neural network has only one hidden layer, we henceforth refer to it as a shallow neural network to distinguish it from more complex architectures such as deep neural networks with numerous hidden layers and LLMs.

### Analyzing model performance

Due to the imbalance of our dataset (N=34, of which nfvPPA = 10), classification ability was primarily assessed by the macro (i.e., unweighted) average of each subtype’s F1 score (aka F1 macro). Each subtype’s F1 score is defined as the harmonic mean of its sensitivity (% of individuals with the subtype being predicted as that subtype) and positive predictive value (% of subtype predictions being correct, PPV).

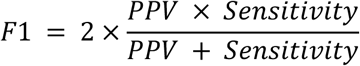

We used the macro-average of each subtype’s F1 score (rather than a weighted average) because we consider both classes (nfvPPA and lv) to be equally important. For each classifier, we also report each subtype’s sensitivity, specificity (% of individuals with the other subtype being predicted as the other subtype, i.e., the other subtype’s sensitivity), PPV, and negative predictive value (NPV, % of the other subtype’s predictions being correct, i.e., the other subtype’s PPV). In sum, our paper’s main evaluation metric is F1 macro, which is defined as the macro (i.e., unweighted) average of each subtype’s F1 score, which is in turn defined as the harmonic mean of a subtype’s PPV and sensitivity.

To evaluate whether a classification algorithm performed above chance, we compared its performance to a naive classifier (the most frequent class baseline), which predicts each sample as the most common class in our dataset (i.e., lvPPA). This baseline was implemented using Scikit-Learn’s DummyClassifier model, with the strategy parameter set to “most_frequent.” In our study, the most frequent class baseline achieves 71% (24/34) accuracy but 41% F1 macro. This is evidence that for unbalanced datasets, F1 macro is a better evaluation metric of classification performance than accuracy. In summary, we evaluate each classifier using the F1 macro metric and determine whether each classifier outperforms the most frequent class baseline.

### Cross-validation and hyperparameter tuning

We adopted leave-one-out nested cross-validation (LOONCV) because it provides an unbiased estimate of true generalization performance for samples of fewer than 100^57^. We used five inner folds for hyperparameter tuning. We used *n* outer folds (*where n* is the number of samples, i.e., leave-one-out) to ensure that the reported results were applicable across all included participants. This was accomplished using Scikit-learn. Hyperparameters were tuned to optimize each classifier’s F1 macro. For each classifier, we employed Receiver Operating Characteristic (ROC) analysis to determine the optimal classification threshold, following the methodology outlined by Bedrick^58^. Optimal thresholds are reported in Supplementary Table 7. To confirm that classification performance was not dependent on this threshold, we evaluated whether the classifier’s balanced accuracy was comparable to the classifier’s area-under-curve score (AUC), which represents the predictive power of a classifier without the need for a specific threshold^20^. This was the case for the best classifier in both the NDL and DL. To better understand AUC scores, we visualized ROC curves, which display the true positive rate and false positive rate of a classifier across different thresholds, with an optimal model having an AUC closer to 1^20^.

To prevent overfitting, we adhered to the N–1 feature-to-sample ratio criterion^18^, which requires that the number of input features be smaller than the number of training samples. When LOOCV is applied to our sample size of 34, each training fold includes 33 samples. Given that we derived 34 features (see Supplementary Table 1), we could not satisfy this criterion by feeding all these features into a classification algorithm. Thus, we trained separate classifiers for each feature set. In summary, we avoided overfitting by ensuring that the number of features used as input to the classifier was significantly smaller than our sample size.

### Feature importance and univariate testing

After building each classifier, we conducted feature permutation analysis^59^, an algorithm-agnostic feature importance approach in contrast to the tree-specific algorithm used in Lukic et al.^18^. For each feature, we calculated the average decrease in classification performance from 10000 random permutations of that feature. Feature permutation analysis was conducted with ScikitLearn’s *permutation_importance* method. It is essential to note that the sum of the individual feature importances may not equal 100% because a naive classifier would not yield 0% classification performance.

After extracting feature importance, univariate testing was conducted to determine if each of the top three features from each feature set differed significantly between nfvPPA and lvPPA. Depending on the data characteristics, we used independent-samples t-tests or Mann-Whitney U tests via the Python package SciPy. In sum, for each feature set’s classifier, we apply feature permutation analysis to identify the three most important features, followed by univariate tests to determine significance.

### Ensemble modeling

We implemented a late-fusion^25^ ensemble model that combined nfvPPA prediction probabilities from each feature set’s classifier as input to the aforementioned classification algorithms. Each feature set’s classifier is defined as the classification algorithm that achieves the highest F1 macro score, following previous PPA classification studies that evaluate multiple classification algorithms and report the performance of the most accurate^17,20,21^. Our ensemble classifier integrates fluency (speech-timing), lexical production (corpora-derived word-level parameters and linguistic features), syntax (linguistic features), and discourse (image–text congruence), in contrast to individual classifiers that consider these abilities in isolation. Like the final classifier for each feature set, the ensemble classifier will be defined as the ML-based classification algorithm that achieves the highest balanced accuracy score given the ensemble’s input. Then, the importance of each feature set to the ensemble classifier was computed using the aforementioned feature importance algorithm.

### Comparing the performance of NDL and DL

We compared the performance of the following classifiers to address whether NDL classifiers outperformed DL classifiers. To do this, we used McNemar tests from the mlxtend package^60^ following Dial et al.^61^. From the predictions of two classifiers compared, a 2x2 contingency table was formed using the *mcnemar_table* function from the mlxtend package. From this contingency table, the McNemar test statistic and corresponding p-value were computed using the *<u>mcnemar</u>* function from the mlextend package. This was also done to compare the classification performance between Spanish and Catalan samples, evaluating whether features are derived robustly in both languages (Supplementary Materials section, Comparing classification performance between Spanish and Catalan).

## Results

### Classification performance for individual feature sets Classification performance in the non-dominant language

Classification performance of individual feature sets using samples in the non-dominant language (NDL: lvPPA n = 24, 18 Spanish, 6 Catalan; nfvPPA = 10; 5 Spanish, 5 Catalan; see Online Methods section *Participants*) revealed the following main findings (Table 2): (1) All feature sets outperformed the most frequent class baseline (i.e., chance) by ≥36% (F1 macro, unweighted average of each subtype’s F1 score, which is in turn defined as the harmonic mean of a subtype’s positive predictive value and sensitivity, see Materials and Methods section *Analyzing model performance*), confirming that each feature set successfully captures differences between the variants. (2) *Word-level parameters* derived from corpora achieved the highest classification performance (F1 macro = 93%, accuracy = 94%). (3) The second-best performing feature set was *Image-text congruence* (F1 macro = 86%, accuracy = 88%). (4) The next-best performing feature set was *speech-timing measures* (F1 macro = 80%, accuracy = 85%). (5) The worst-performing feature set was the *linguistic feature set* derived with NLP tools excluding corpora (F1 macro = 73%, accuracy = 76%). (6) All feature sets achieved better classification performance in lvPPA (F1’s 83-92% across feature sets) compared to those with nfvPPA (F1’s 64-80%).

**Table 2:** Percent classification performance in the non-dominant language by feature set. Only the performance for the best (highest F1 macro) classification algorithm is reported here. To see performance for all classification algorithms, please see Supplementary Table 8. A subtype’s sensitivity is the % of patients with that subtype who are correctly predicted as that subtype. A subtype’s positive predictive value (PPV) is the % of subtype predictions that are correct. A subtype’s F1 is the harmonic mean of its sensitivity and PPV. A subtype’s specificity is the % of patients with the other subtype being predicted as the other subtype (i.e., the other subtype’s sensitivity). A subtype’s negative predictive value (NPV) is the % of the other subtype’s predictions being accurate (i.e., the other subtype’s PPV). F1 macro is the unweighted (macro) average of each subtype’s F1. AUC = Area under the curve. Evaluation metrics are further discussed in the Online Methods section *Analyzing model performance*.

|  |  | Feature set |  |  |  |
| --- | --- | --- | --- | --- | --- |
|  |  | Image-Text congruence | Speech-timing | Word-level | Linguistic |
| Best classification algorithm |  | Decision Tree | Decision Tree | Support Vector Machine | Decision Tree |
| nfvPPA | sensitivity | 80 | 60 | 90 | 70 |
|  | PPV | 80 | 86 | 90 | 58 |
|  | F1 | 80 | 71 | 90 | 64 |
|  | specificity | 92 | 96 | 96 | 79 |
|  | NPV | 92 | 85 | 96 | 86 |
| lvPPA | sensitivity | 92 | 96 | 96 | 79 |
|  | PPV | 92 | 85 | 96 | 86 |
|  | F1 | 92 | 90 | 96 | 83 |
|  | specificity | 80 | 60 | 90 | 70 |
|  | NPV | 80 | 86 | 90 | 58 |
| F1 macro |  | 86 | 80 | 93 | 73 |
| Accuracy |  | 88 | 85 | 94 | 76 |
| AUC |  | 76 | 78 | 88 | 71 |

In the NDL, we report the three most important features, determined via permutation analysis, for each feature set’s classifier as follows. The three most important *word-level parameters* included the following: concreteness (34% drop in classification performance via permutation analysis, see Online Methods section *Feature importance and univariate testing*), which was significantly larger in nfvPPA compared to lvPPA (Supplementary Table 9, see Methods section *Feature importance and univariate testing)*; age of acquisition (18%), which did not differ significantly between the PPA variants; verb frequency (12%), which also did not differ significantly between the PPA variants (Supplementary Table 9).

The three most important speech-timing measures included the following: articulation rate (34%), which was significantly faster in lvPPA compared to nfvPPA; number of words per minute (17%), which was significantly faster in lvPPA compared to nfvPPA; number of propositions per minute (4%), which was significantly faster in lvPPA compared to nfvPPA (Supplementary Table 10).

The three most important linguistic features included the following: total number of words (33%), which was significantly larger in lvPPA compared to nfvPPA; average sentence length (14%), which did not differ between the variants, and propositional density (12%), which was significantly larger in lvPPA compared to nfvPPA (Supplementary Table 11).

### Classification performance in the dominant language

Classification performance of individual feature sets using samples in the dominant language (DL: lvPPA n = 24, 6 Spanish, 18 Catalan; nfvPPA n = 10, 5 Spanish, 5 Catalan; see Online Methods section *Participants*) revealed the following key findings (Table 3): (1) All feature sets outperformed the most frequent class baseline (i.e., chance) by ≥ 40% (F1 macro), confirming that each feature set in the DL captures differences between the variants. (2) *Linguistic features* achieved the highest classification performance (F1 macro = 87%, accuracy = 88%), nearly equal to (3) the second-best performing feature set, *word-level parameters* (F1 macro = 86%, accuracy = 88%). (4) The next-best performing feature set was *Image-text congruence scores* (F1 macro = 82%, accuracy = 85%). (5) Lastly, our worst-performing feature set in the DL was *speech-timing* measures (F1 macro = 70%, accuracy = 79%). (6) All feature sets achieved better classification performance of individuals with lvPPA (F1’s 70%-92%) compared to those with nfvPPA (F1’s 64%-83%).

**Table 3:** Percent classification performance in the dominant language by feature set. Only the performance for the best (highest F1 macro) classification algorithm is reported here. To see performance for all classification algorithms, please see Supplementary Table 12. A subtype’s sensitivity is the % of patients with that subtype who are correctly predicted as that subtype. A subtype’s positive predictive value (PPV) is the % of subtype predictions being correct. A subtype’s F1 is the harmonic mean of its sensitivity and PPV. A subtype’s specificity is the % of patients with the other subtype being predicted as the other subtype (i.e., the other subtype’s sensitivity). A subtype’s negative predictive value (NPV) is the % of the other subtype’s predictions being accurate (i.e., the other subtype’s PPV). F1 macro is the unweighted (macro) average of each subtype’s F1. AUC = Area under the curve. Evaluation metrics are further discussed in the Online Methods section *Analyzing model performance*.

|  |  | Feature set |  |  |  |
| --- | --- | --- | --- | --- | --- |
|  |  | Image-Text Congruence | Speech-timing | Word-level | Linguistic |
| Best classification algorithm |  | Shallow Neural Net | Shallow Neural Net | Decision Tree | Gradient Boosting |
| nfvPPA | sensitivity | 70 | 90 | 80 | 100 |
|  | PPV | 78 | 50 | 80 | 71 |
|  | F1 | 74 | 64 | 80 | 83 |
|  | specificity | 92 | 62 | 92 | 83 |
|  | NPV | 88 | 94 | 92 | 100 |
| lvPPA | sensitivity | 92 | 64 | 92 | 83 |
|  | PPV | 88 | 94 | 92 | 100 |
|  | F1 | 90 | 70 | 92 | 91 |
|  | specificity | 70 | 90 | 80 | 100 |
|  | NPV | 78 | 50 | 80 | 71 |
| F1 macro |  | 82 | 70 | 86 | 87 |
| Accuracy |  | 85 | 71 | 88 | 88 |
| AUC |  | 76 | 81 | 73 | 90 |

In the DL, we report the three most important features for each feature set’s classifier as follows. The three most important *word-level parameters* included the following: concreteness (32%), which was significantly larger in nfvPPA compared to lvPPA; word length by phonemes (8%), which did not differ significantly between the PPA variants; verb frequency per million (2%), which did not differ significantly between the PPA variants (Supplementary Table 9).

The three most important *speech-timing measures* included the following: articulation rate (number of syllables divided by phonation time, reflecting motor speech function more purely than speech rate which includes pauses between words, 18% via permutation analysis), which was significantly faster in lvPPA compared to nfvPPA; number of pauses (11%), which did not differ significantly between the PPA variants; and speech rate (number of syllables per second, 11%), which did not differ significantly between the PPA variants (Supplementary Table 10).

The three most important *linguistic features* included the following: subordination index (32%), which was significantly larger in lvPPA compared to nfvPPA; open-closed words ratio (12%), and part-word repetitions (5%), both of which did not differ significantly between the PPA variants (Supplementary Table 11).

### Ensemble classification performance

While individual feature sets demonstrated strong diagnostic potential, an ensemble model combining each feature set’s individual classifier was built to determine whether a more comprehensive classifier could achieve the robust performance required for clinical application. To build an ensemble model, we derived lvPPA prediction probabilities from the four feature sets’ classifiers for each patient, which were then used as input to classification algorithms (late fusion^24^; Figure 1; see Methods section *Ensemble modeling*). We built two ensemble models, one using responses in the NDL and another using responses in the DL.

**Figure 1.**
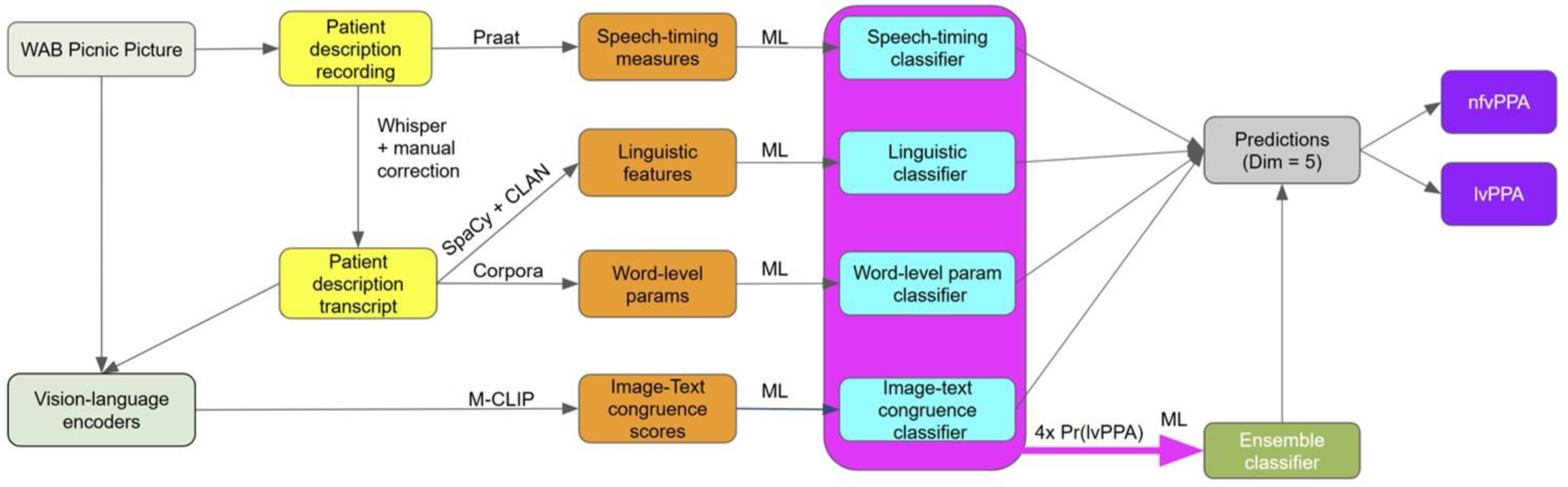
Workflow to differentiate nfvPPA from lvPPA using connected speech. *Note.* Summary of feature extraction workflow starting from picture description recordings. First, *speech-timing measures* are derived from the recording with Praat. Then, the recording is transcribed with Whisper, which is then corrected manually. From the transcription, *linguistic features* are derived with SpaCy and CLAN, and *word-level parameters* are derived from corpora. *Image-text congruence scores* are derived by feeding the transcript and picture into vision-language encoders (i.e., Multilingual-CLIP). Machine learning is used to create a classifier for each feature set. Each classifier’s lvPPA prediction probability is then used to build a multimodal ensemble classifier, which predicts whether the individual has nfvPPA or lvPPA.

### Non-dominant language (NDL) ensemble classification performance

Our NDL ensemble achieved excellent classification performance (F1 macro = 86%, accuracy = 88%, AUC = 79%; Table 4), but underperformed the NDL *word-level parameters classifier* by 7% F1 macro. The ensemble misclassified two individuals with nfvPPA and two individuals with lvPPA (Figure 2). *Word-level parameters* contributed most to the NDL ensemble (41%, permutation analysis), followed by *image-text congruence scores* (5%), *speech-timing measures* (0%), and *linguistic measures* (0%).

**Figure 2.**
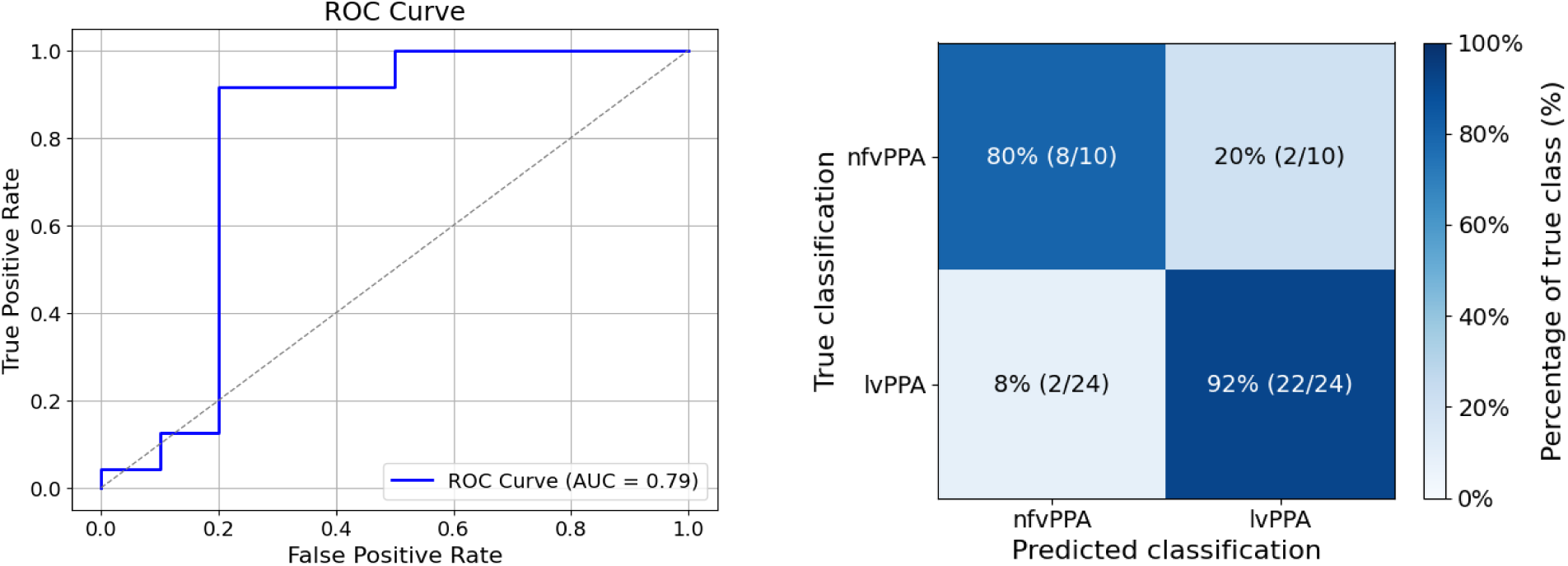
Visualizing performance of the NDL ensemble classifier. *Note.* Left-hand side: Receiver operating characteristic (ROC) curve with the mean area under the curve (AUC). Right-hand side: confusion matrix

**Table 4:** Percent classification performance of the ensemble model for the non-dominant language (NDL) and the dominant language (DL). PPV = Positive Predictive Value, NPV = Negative Predictive Value. Evaluation metrics are defined in the Methods section, *Analyzing model performance*.

|  |  | Classification performance (%) |  |
| --- | --- | --- | --- |
|  |  | NDL | DL |
| Best classification algorithm |  | Gradient Boosting |  |
| nfvPPA | sensitivity | 80 | 80 |
|  | PPV | 80 | 100 |
|  | F1 | 80 | 89 |
|  | specificity | 92 | 100 |
|  | NPV | 92 | 92 |
| lvPPA | sensitivity | 92 | 100 |
|  | PPV | 92 | 92 |
|  | F1 | 92 | 96 |
|  | specificity | 80 | 80 |
|  | NPV | 80 | 100 |
| F1 macro |  | 86 | 92 |
| Accuracy |  | 88 | 94 |
| AUC |  | 79 | 95 |

### Dominant-language (DL) ensemble classification performance

Our DL ensemble achieved better classification performance than its NDL counterpart (F1 macro = 92%, accuracy = 94%, AUC = 95%; Table 4), outperforming each feature set’s DL classifier by ≥ 5% F1 macro. The ensemble misclassified two individuals with nfvPPA (Figure 3) *Word-level parameters* contributed most to the DL ensemble (34%), followed by *linguistic features* (29%), *image-text congruence scores* (0%), and *speech-timing measures* (0%).

**Figure 3.**
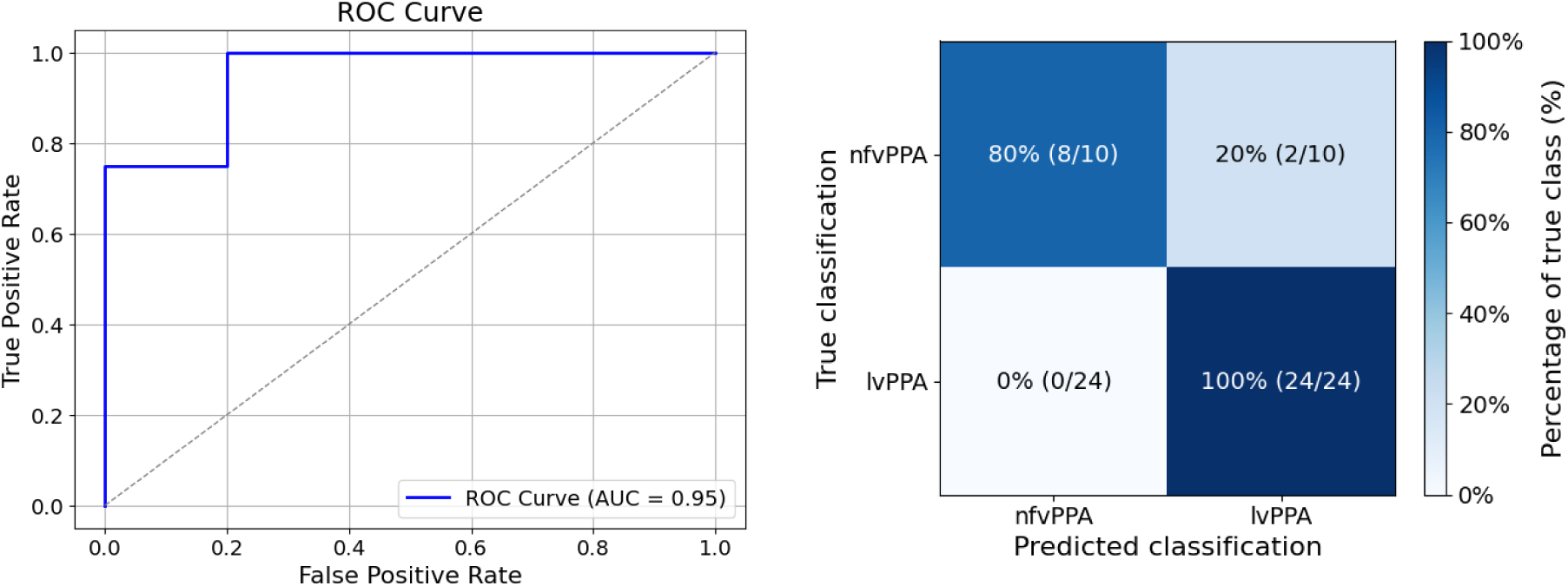
Visualizing performance of the DL ensemble classifier. *Note.* Left-hand side: Receiver operating characteristic (ROC) curve with the mean area under the curve (AUC). Right-hand side: confusion matrix

### Comparing classification performance between individual feature sets and ensemble modeling in non-dominant and dominant languages

No significant differences in classification performance emerged between the NDL and DL across individual feature sets or the ensemble (Table 5). Numerically, the NDL outperformed the DL for *word-level parameters* (7%, *p* = 0.63), *image-text congruence scores* (4%, *p* = 0.63), and *speech-timing measures* (10%, *p* = 0.58, McNemar’s test, see Online Methods section *Comparing the performance of NDL and DL*). The DL numerically outperformed the NDL for *linguistic features* (14%, *p* = 0.34), and the ensemble (6% F1 macro, *p* = 0.63). The best NDL classifier (*word-level parameters*) performed nearly equivalently to the best DL classifier (ensemble) by 1% (p = 1). In sum, classification performance was not significantly influenced by language dominance.

**Table 5:** Comparing percent F1 macro classification performance between the non-dominant and dominant language for each feature set and the ensemble model. The best classifiers in the non-dominant and dominant languages are bolded.

| Feature set | Non-dominant language (most discriminative features/feature sets) | Dominant language (most discriminative features/feature sets) | McNemar p |
| --- | --- | --- | --- |
| Ensemble | 86 (Word-level parameters, image-text congruence) | **92** (Word-level parameters, Linguistic features) | 0.63 |
| Image-text congruence | **86** | 82 | 1 |
| Speech timing measures | **80** (Articulation rate, # of words per minute, # propositions per minute) | 70 (Articulation rate) | 0.58 |
| Word-level parameters | **93** (Concreteness) | 86 (Concreteness) | 0.63 |
| Linguistic | 73 (# of words, propositional density) | **87** (Subordination) | 0.34 |
| Best performing model (word-level for NDL and ensemble for DL) | **93** | **92** | 1 |

## Discussion

The current study evaluated whether multilingual multimodal machine learning (M^3^L) applied to brief connected speech samples (i.e., picnic scene descriptions from the Western Aphasia Battery) can effectively differentiate nfvPPA and lvPPA in Catalan-Spanish bilinguals. Overall, our findings demonstrate that this M^3^L-based approach reliably distinguishes PPA variants in both their dominant (DL) and non-dominant languages (NDL), and that features previously identified as discriminative in monolingual speakers retained their diagnostic value in our bilingual sample.

We first evaluated classification performance by language dominance. All feature sets (i.e., *word-level parameters, image-text congruence scores, speech-timing measures, linguistic features*) outperformed chance, regardless of dominance status. Also, across all feature sets, NDL performance was comparable to that of DL. This finding suggests that accurate classification does not require testing in the dominant language, despite clinicians’ tendency to default to it^10^.

Second, we examined whether ensemble modeling improved performance. In the DL, the ensemble provided only a slight improvement over the most discriminative feature set (*linguistic features*). In the NDL, the ensemble underperformed the best feature set (*word-level parameters*). Taken together, these results indicate that ensemble modeling did not improve classification accuracy in either language and that select feature sets alone may be sufficient for accurate classification, particularly in the NDL.

Third, we assessed whether the most discriminative features aligned with prior PPA research. The features driving classification have previously been identified in monolingual English and Spanish PPA classification studies^3,17,19,21,39,49,50,62–64^. Thus, our findings extend prior work by demonstrating that these markers retain discriminative value in bilingual speakers, a population that is particularly challenging to classify accurately due to natural features of bilingual speech and language^7^.

Feature set contributions to the ensemble differed by language dominance. Although both the NDL and DL ensemble classifiers were primarily driven by word-level features, particularly concreteness, the relative contributions of other feature sets differed by language dominance. In the NDL group, image–text congruence contributed more strongly to the ensemble, whereas in the DL group, linguistic features, and speech-timing measures, particularly subordination index and articulation rate, played a larger role. This divergence likely reflects how effectively individual measures distinguished variants within each group; features with stronger variant-related differences provided more diagnostic information, thereby carrying greater weight in overall classification performance. For example, the subordination index differed significantly between variants only in the DL group (Supplementary Table 11), which may help explain the greater contribution of linguistic features to the DL ensemble. This interpretation is broadly consistent with previous studies reporting that measures of subordination distinguish PPA variants in monolingual English speakers^18,64^. Conversely, image–text congruence showed stronger variant-related differences in the NDL group than in the DL group (Supplementary Table 13; Table 5), which may account for its greater contribution to the NDL ensemble. Together, these findings show that although ensemble classification performance was comparably accurate across language-dominance groups, the relative importance of specific sources of information may vary as a function of language dominance.

In the paragraphs that follow, we comment on the most discriminative features by each feature set that met the following conditions: 1) they emerged as one of the three most important features and 2) significantly differed between variants. The only *word-level parameter* that fulfilled both conditions was concreteness, which was significantly greater in nfvPPA compared to lvPPA in both the NDL and DL. This implies that individuals with nfvPPA are more likely to produce words directly and concisely referring to a perceptible entity (e.g., ‘sailboat’), whereas those with lvPPA are more likely to use circumlocutions to describe objects that they can’t name directly (e.g., ‘this is a vehicle that moves people on water’). This finding aligns with previous research indicating that monolingual English and Spanish speakers with nfvPPA maximize communicative efficiency by using relatively preserved lexical-semantic processing in the context of reduced syntactic complexity and speech production difficulties^40,65^.

The most discriminative *linguistic features* were the total number of words produced, the subordination index, and propositional density. First, the total number of words produced was significantly lower in nfvPPA than in lvPPA for both the NDL and the DL, aligning with previous monolingual PPA studies in English^62^ and Spanish^21,63^, and may be influenced by reduced syntactic complexity and/or motor speech impairments characteristic of nfvPPA. Second, the subordination index was significantly higher in lvPPA compared to nfvPPA (in the DL only), consistent with prior findings in monolingual English speakers with PPA. This suggests that individuals with nfvPPA experience broader difficulty with syntactic complexity, particularly hierarchical structures involved in subordination^18,64^. However, this does not align with a previous study of Spanish-speaking individuals with PPA^21^, which found that their subordination index did not differentiate nfvPPA from lvPPA. This perceived discrepancy may stem from differences in how the subordination index was defined: their simpler measure (number of grammatical clauses divided by number of grammatical sentences) captures clause-internal indirect verb relations, whereas our index counts subordinate clauses containing at least one finite matrix verb.

Third, propositional density (also referred to as idea density, defined as the percentage of words that can be combined with nouns to form propositions i.e., verbs, adjectives, adverbs, prepositions and conjunctions) was also significantly lower in nfvPPA than in lvPPA for both the NDL and the DL, consistent with prior work in monolingual English speakers with PPA^39^. We interpret this as reflecting a reduced ability in individuals with nfvPPA to combine nouns with relational words that form propositions. Consistent with this interpretation, participants with nfvPPA produced significantly fewer adverbs (aligning with Graham et al.^66^; Supplementary Table 14) and fewer subordinating conjunctions. Because subordinating conjunctions support clause embedding and the expression of relationships between ideas, their reduced use likely contributed directly to the lower propositional density observed in this group.

The most discriminative *speech-timing measures* were articulation rate (number of syllables divided by phonation time), number of words per minute, and number of propositions per minute. Each reflects the speed of spoken production across syllables, words, and propositions. First, in both the NDL and DL, articulation rate was significantly slower in nfvPPA than in lvPPA. This aligns with previous findings in monolingual English speakers with PPA^20,50^, which attribute slower articulation rates to the motor speech impairments that often accompany nfvPPA. Second, in the NDL only, the number of words per minute was significantly lower in nfvPPA compared to lvPPA, aligning with previous studies in monolingual Spanish speakers with PPA^21^ and monolingual English speakers with PPA^49^. A possible explanation for this NDL-specific finding is that speech rate was lower in the NDL than in the DL for both variants (Supplementary Table 10), suggesting the NDL is more susceptible to slowing of speech rate. Third, in both the NDL and the DL, the number of propositions per minute was significantly lower in nfvPPA than in lvPPA, consistent with previous studies indicating that both the number of propositions and speech rate were significantly lower in nfvPPA than in lvPPA^39,49^.

Considering our results more holistically (i.e., across feature sets), the established variant-specific speech-language impairments were reflected in the most discriminative features that emerged. We observed evidence of greater word-retrieval difficulties (i.e., lower concreteness and image-text congruence, Supplementary Table 9 and Supplementary Table 13) in lvPPA compared to nfvPPA, and greater difficulties with grammar (e.g., lower subordination index) and motor speech impairment (e.g., articulation rate, number of words per minute) in nfvPPA compared to lvPPA. In sum, the most discriminative features were consistent with known spared and impaired speech-language features associated with each respective variant^1^. Taken together, these findings indicate that the features that best differentiate nfvPPA and lvPPA are broadly similar across bilingual and monolingual populations, although further work will be needed to confirm the generalizability of this pattern while accounting for language-specific features.

### Limitations

A limitation of our sample was unbalanced sample size between PPA variants, which could have driven the pattern of consistently higher F1 scores for lvPPA (as they had a larger sample size). Therefore, future work should evaluate the approach described herein with a more balanced and larger dataset.

A second limitation of our study is that its immediate clinical applicability is limited by its reliance on human transcription. That being said, most features used in the study can be automatically derived from utterance-segmented text files. This is not the case for the two linguistic features of retracing and part-word repetitions, which can only be derived from transcriptions formatted specifically for CLAN. These limitations evidently do not apply to the speech-timing measures, which can be automatically derived from audio files but did not achieve optimal classification performance (i.e., ≤ 85% F1 macro). An important next step will be to evaluate whether similar classification performance can be achieved using automated transcriptions generated with Whisper^35^. If successful, this would yield a fully automated 1–2-minute assessment that is significantly shorter than current clinical assessments (which typically last multiple hours). This would potentially address inequities in PPA differential diagnosis and reduce the workload of bilingual clinicians.

## Conclusions

Although most of the world’s population is bilingual, methods for differentiating PPA variants in bilingual speakers remain limited. Building on recent advances in multilingual, multimodal machine-learning (M^3^L), the present study demonstrates that Catalan-Spanish bilingual individuals with nfvPPA and lvPPA can be differentiated with high accuracy using a largely automated, time-efficient (1–2 minutes), and ecologically valid connected-speech–based method. To our knowledge, this is the first study to examine variant classification in bilingual individuals with PPA, the third in a non-English context, and the first to evaluate the impact of language dominance on classification performance. Importantly, ensemble classification performance did not differ by language dominance, but the underlying feature set contributions to the ensemble did. Given the linguistic diversity of individuals affected by PPA, the development of assessment approaches that are both accessible and equitable is critical. The reported findings represent a meaningful step toward more inclusive diagnostic tools for multilingual populations.

## List of abbreviations

M³L: Multilingual Multimodal Machine Learning
nfvPPA: nonfluent variant primary progressive aphasia
lvPPA: logopenic variant primary progressive aphasia
PPA: primary progressive aphasia
ML: machine learning
M-CLIP: Multilingual Contrastive Learning Image Pretraining
svPPA: semantic variant primary progressive aphasia
TDP-43-C: transactive response DNA-binding protein 43 type C pathology
N: number (i.e., sample size)
NLP: Natural Language Processing
NDL: non-dominant language
DL: dominant language
CSF: cerebrospinal fluid
MMSE: Mini-mental state examination
WAB-R: Western Aphasia Battery-Revised
rVAD: Robust voice activity detection method
CLAN: Computerized Language Analysis
NPV: Negative Predictive Value
PPV: Positive Predictive Value
LOONCV: leave-one-out nested cross-validation
ROC: Receiver Operating Characteristic
AUC: Area under curve
L1: First language
L2: Second language

## Declarations

### Ethics approval and consent to participate

The study was approved by the ethics committees of Hospital de Sant Pau and Hospital Clínic de Barcelona, as well as the Institutional Review Board at the University of Texas at Austin. All participants provided written informed consent.

### Consent for publication

Not applicable

### Availability of data and materials

The data supporting the findings of this study are subject to HIPAA regulations. However, this data may be made available upon request and with the execution of appropriate data use agreements. Please contact the corresponding author, Miguel Ángel Santos Santos, for data use inquiries at.

### Author’s contributions

L.S.P: Conceptualization, Data Curation, Formal Analysis, Investigation, Methodology, Project Administration, Software, Writing - original draft, Writing - review & editing

A.P.C: Data Curation, Investigation, Methodology, Software, Validation, Writing - original draft, Writing - review & editing

N.M.C. Conceptualization, Investigation, Methodology, Software, Validation, Writing - review & editing

S.K.M., C.W.R., J.C.H.C, J.F.M: Investigation, Methodology, Data Curation, Software, Validation, Writing - review & editing

W.A.M Data Curation, Writing - review & editing

J.J.L: Conceptualization, Supervision, Writing - Review & editing

F.L.: Data Curation, Methodology, Validation, Writing - review & editing

N.Z., S.R, I.I, S.B, A.Llado, J.F, A.Leo, R.S: Data Curation, Writing - review & editing

M.L.H: Conceptualization, Methodology, Writing - review & editing

M.A.S.S: Conceptualization, Data Curation, Funding Acquisition, Investigation, Methodology, Project Administration, Resources, Software, Supervision, Writing - review & editing

S.M.G: Conceptualization, Data Curation, Funding Acquisition, Investigation, Methodology, Project Administration, Resources, Software, Supervision, Validation, Writing - original draft, Writing - review & editing

## Acknowledgements

We thank the Basque Center on Cognition, Brain and Language (BCBL) for access to their internal ESPAL database. L.S.P. would like to thank Eric Wuesthoff for their support with science communication.

