## Supplementary Materials for "Differentiating nonfluent/agrammatic and logopenic primary progressive aphasia in Catalan-Spanish bilinguals by applying multilingual multimodal machine learning (M³L) to connected speech"

Supplementary Table 1: Definitions and sources for each feature in each derived feature set

| Feature set | Specific Feature name | Feature definition | Source |
|---|---|---|---|
| Linguistic | Total number of words | Excludes fillers and interjections, listed below | SpaCy[37] |
| | Proportion of fillers | # of fillers / # of words<br><br>List of fillers in both languages: ahm, uff, ai, ei, vale, emh, ehm, pues, bueno, clar, doncs, ba, ah, er, hm, mm, uh, eh, em*, ja, ay<br><br>*em is not a filler in Catalan as it’s a clitic/pronoun | SpaCy |
| | Type-token ratio | # of unique words / # of words, measure of lexical diversity | SpaCy |
| | Noun-to-verb ratio | # of nouns / # of verbs | SpaCy |
| | Propositional density | Percentage of words labeled with the following part of speech tags: verbs, adjectives, adverbs, prepositions, and conjunctions / number of words | SpaCy |
| | Open-to -closed class words ratio | # of words with open class POS tags (verbs, nouns adjectives, nouns) / # of words with closed class POS tags (defined as non-open class POS tags) | SpaCy |
| | Average sentence length | # of words / # of sentences. # of sentences computed by summing # of periods, question marks and exclamation points. | SpaCy |
| | Subordination index | # of complementizers / # of sentences<br><br>Complementizers: words tagged as SCONJ before the last finite verb or relative pronouns (only counted if a sentence has a finite verb)<br><br>Exception rule 1: Como for Spanish, com for Catalan if como is followed immediately by an SCONJ, then como is considered to be an SCONJ<br><br>Exception rule 2: ‘donde’ tagged as a pronoun is a complementizer<br><br>From feature dictionary: Number of subordinate clauses that include at least one finite matrix verb | SpaCy |
| | retracings | # of retracings / # of words | CLAN[36] |

| | | | |
|---|---|---|---|
| | whole-word repetitions | # of whole-word repetitions / # of words | SpaCy |
| | part-word repetitions | # of part-word repetitions / # of words | CLAN |
| Corpora-derived word-level parameters | Word frequency per million | frequency per million, averaged across each word with an available rating* | BCBL's Espal[43] for Spanish, SUBTLEX-CAT[42] for Catalan |
| | Verb frequency per million | frequency per million, averaged across each verb with an available rating* | Above |
| | Noun frequency per million | frequency per million, averaged across each noun with an available rating* | Above |
| | % high frequency verbs | % of verbs whose lemmas are in the top 10 most frequent verb lemmas, as defined below<br><br>Spanish's top 10 most frequent verb lemmas: ser, haber, tener, estar, hacer, poder, decir, dar, ver, ir<br><br>Catalan's top 10 most frequent verb lemmas: ser, fer, anar, tenir, haver, estar, voler, veure, passar, deixar | Above |
| | word length by syllables | # of syllables in a word, averaged across each word where a value is available* | Above |
| | word length by phonemes | Number of phonemes in a word, averaged across each word. | Espal[43] for Spanish, CharsiuG2P[46] for Catalan |
| | concreteness | Rating of degree to which a concept denoted by a word refers to a perceptible entity, averaged across each word where a rating is available* | Thompson et al.[45] |
| | age of acquisition | Approximate minimum age when a word is learned, averaged across each word where rating is availability* | Alonso et al.[41] for Spanish, Luniewska et al.[44] for Catalan |
| Speech-Timing measures | Speech rate | # syllables / # seconds | Parselmouth[48] script in Feinberg et al.[47] |
| | Average syllable duration | Phonation (aka speaking) time / # syllables | |
| | Articulation rate | # syllables / phonation time | |
| | Speech-pause ratio | speaking time / non-speaking time | |

| | | | |
|---|---|---|---|
| | # of pauses | Number of pauses | |
| | mean pause duration | An additional line of code was written to save the text grid object in Feinberg's original script, which onsists of time intervals for each silent and non-silent period, from which the mean and variance of pause duration are computed. | Extending the Parselmouth[48] script from Feinberg et al[47] |
| | variance of pause duration | | |
| | # words/minute | # of words is derived with SpaCy. Duration of audio file derived with Feinberg's script. | SpaCy, Feinberg's script |
| | # propositions /minute | # of propositions is derived with SpaCy. Duration of audio file derived with Feinberg's script | SpaCy, Feinberg's script |
| Image-text congruence | | A measure of the semantic congruence between a text and a picture according to a vision-language encoder[23]; better picture descriptions will have higher image-text congruence scores. 4 image-text congruence scores were derived using the 4 vision-language encoders of Multilingual CLIP[54] | Multilingual(M) -CLIP[54] |

*If a word does not have an available value, the rating for the word's lemma is used if available instead

**Validation of linguistic and speech-timing feature extraction**

Compared to English, NLP tools for deriving linguistic features in Spanish and Catalan are lacking and thus require extra post-processing and validation. For SpaCy's POS tagger, morphologizer, and tokenizer, in Spanish and Catalan, we added several additional rules to ensure their error rate was negligible (Supplementary Tables 2-6). A group of linguists and speech-language pathologists with a strong background in romance languages manually validated the derived linguistic features for a randomized subset of samples in Catalan and Spanish

Measures related to speech timing were extracted from the audio samples using Praat, as described in Supplementary Table 1. The output was validated for a randomized subset of samples in Catalan and Spanish through the following steps: 1) ensuring a sufficient signal-to-noise ratio (20-40 dB); 2) ensuring the error rate for computing the number of syllables <10%; 3) ensuring the labels of "silent" and "sounding" in the Praat textgrid accurately matched the audio segments.

**Finetuning the Catalan SpaCy model**

To improve the accuracy of SpaCy's Catalan part-of-speech (POS) tagger, we finetuned its model[1] on 14 transcriptions from Catalan-speaking PPA patients who were not used in this paper. From this, we added tokenization rules shown in Supplementary Table 2. Supplementary Table 3 presents POS tagging rules for specific words that are universally applied in all contexts. Rules from both Supplementary Tables 2 and 3 were added to the model's attribute ruler, meaning the model incorporated them. This functionality is available in SpaCy v3.6. Fine-tuning the model's POS tagger impacted its morphological output, as morphological processing occurs after POS tagging. An implemented post-processing (meaning the SpaCy model itself didn't learn this) rule was that the POS tag for the word 'la' is DET if followed by a noun. A preprocessing rule implemented is that all apostrophes were replaced with a space.

[1]https://spacy.io/models/ca#ca_core_news_trfpart-of-speech

Supplementary Table 2. Catalan Tokenizer rules:

| **String** | **Desired Tokenization** |
|---|---|
| de pressa | depressa* |
| de seguida | deseguida* |
| al | al |
| als | als |
| dels | dels |
| pels | pels |
| cal | cal |
| almar | al mar |
| ambun | amb un |
| hihandues | hi han dues |
| Ihandues | i han dues |
| iel | i el |
| ia | i a |
| iun | i un |
| quequedés | que quedés |
| afora | a fora |
| iaixò | i això |
| neudona | neu dona |

*Included to ensure this word is tagged as an adverb

Supplementary Table 3. Catalan Part-of-speech (POS) tagger rules

| POS tag | Words |
|---|---|
| INTJ | Ahm, uff, ai, ei, vale, emh, ehm, pues, bueno, clar, doncs, ba, ah, er, hm, mm, uh, eh, ja, ay |
| ADV | Aviat, demés, davant, gaire, com, fora, darrere, com, allavorens, mentres, llavons, depressa, deseguida* (see note above about pre-processing), molta |
| DET | Uns, un, unas, l, els, una, altre, aquestes, aquesta, aquest, la, les, tal |
| NOUN | Sopar, mans, manos, castells, chica, chico, filosofia, química, universitat, etcètera, laboratoris, matemàtiques |
| PRONOUN | meus, es, m, algo |
| ADJ | Mateixa, casats, mig, cada |
| VERB | Ajuda, vec, sigut, tenia, ser, és, són, eren, era, estar, estat, caminando, fos, mira, sigui |
| AUX | se |
| ADP | a |
| NUM | seixanta |

Supplementary Table 4. Part-of-speech (POS) and morphologizer (MORPH) error rates of the finetuned Catalan SpaCy model

| | | **# of errors** | |
|---|---|---|---|
| **Sample ID** | **# words** | **POS** | **MORPH** |
| BISE010_PreTx_Catalan_CatRescue | 56 | 3 | 1 |
| BISE010_PreTx_Catalan_ImportantEvent | 14 | 0 | 0 |
| BISE010_PreTx_Catalan_WABPicnic | 89 | 3 | 4 |
| BISE012_PreTx_Catalan_CatRescue | 97 | 1 | 2 |
| BISE012_PreTx_Catalan_ImportantEvent | 233 | 4 | 4 |
| BISE012_PreTx_Catalan_WABPicnic | 131 | 0 | 3 |
| BISD007_PreTx_Catalan_CatRescue | 76 | 3 | 2 |
| BILP017_PreTx_Catalan_WABPicnic | 167 | 9 | 8 |
| BILP015_PostTx_Catalan_PicnicScene | 164 | 4 | 3 |
| BILP014_PreTx_Catalan_ImportantEvent | 160 | 10 | 10 |
| BILP013_PreTx_Catalan_CatRescue | 213 | 8 | 7 |
| BLP011_PreTx_Catalan_CatRescue | 84 | 1 | 1 |
| BILP010_PreTx_Catalan_CatRescue | 171 | 10 | 2 |
| BILP006_PreTx_Catalan_WABPicnic | 252 | 17 | 11 |
| BILP006_PreTx_Catalan_ImportantEvent | 126 | 3 | 4 |
| Sum | 2033 | 76 | 62 |
| **% error rate** | | **3.7** | **3.05** |

As shown in Supplementary Table 4, we evaluated our fine-tuned Catalan SpaCy transformer model on 15 unseen transcriptions (not used to build the classifier or fine-tune the SpaCy model). Across 2033 words in these 15 transcriptions, the POS tagger and the morphologizer were only incorrect for 76 words and 82 words, respectively, meaning the POS and MORPH error rates were 3.7% and 3.05%, respectively.

**Finetuning the Spanish SpaCy model**

Similar to Catalan, we also finetuned SpaCy's Spanish transformer model[2]. The initial POS tagging performance was significantly better than that of Catalan, so we only needed to fine-tune this model on eight Spanish transcriptions. The additional rules are listed in Supplementary Table 5.

Supplementary Table 5. Spanish POS tagger rules

| POS tag | Words |
|---|---|
| INTJ | Ahm, uff, ai, ei, vale, emh, ehm, pues, bueno, clar, doncs, ba, ah, er, hm, mm, uh, eh, em, ja, ay |
| ADV | Sí, cómo |
| NOUN | lao |
| PRONOUN | desto |
| ADJ | Su, sus |

[2]https://spacy.io/models/es#es_dep_news_trf

Supplementary Table 6. Part-of-speech (POS) and morphologizer (MORPH) error rates of finetuned Spanish SpaCy model

| Sample ID | # words | # POS errors | # MORPH errors |
|---|---|---|---|
| BILP006_PreTx_Span_CatRescue | 251 | 6 | 5 |
| BILP006_PreTx_Span_Important_ Event | 223 | 2 | 7 |
| BILP006_PreTx_Span_WABPicnic | 314 | 7 | 6 |
| BILP008_PreTx_Spa_CatRescue | 181 | 1 | 3 |
| BILP009_PreTx_Spa_CatRescue. | 96 | 1 | 3 |
| BILP010_PreTx_Spa_CatRescue | 202 | 1 | 15 |
| BILP010_PreTx_Spa_Important Event | 389 | 2 | 28 |
| BILP010_PreTx_Spa_WABPicnic | 141 | 1 | 3 |
| BILP011_PreTx_Spa_WABPicnic | 116 | 0 | 3 |
| BILP013_PreTx_Spa_WABPicnic | 361 | 3 | 6 |
| BILP015_PostTx_Spa_CatRescue | 198 | 3 | 1 |
| BILP015_PostTx_Spa_WABPicnic | 171 | 7 | 0 |
| BILP017_PreTx_Spa_WABPicnic | 56 | 0 | 1 |
| BILP018_PreTx_Spa_WABPicnic | 255 | 2 | 4 |
| Sum | 2954 | 36 | 85 |

| | | | |
|---|---|---|---|
| % error rate | | 1.2 | 2.9 |

As shown in Supplementary Table 6, we evaluated our fine-tuned Spanish SpaCy transformer model on 15 unseen transcriptions (not used to build the classifier or fine-tune the SpaCy model). Across 2954 words in these 15 transcriptions, the POS tagger was incorrect for 25 words, meaning its word error rate was 1.2%, and the morpoholologizer was incorrect for 26 words, meaning its word error rate was 2.8%.

**Lemmatization evaluation and correction**

We observed that SpaCy's lemmatizer for both languages was very accurate. We implemented only one fix: the lemma for "sé" is "saber", not "ser" as initially indicated by SpaCy.

**Optimal classification threshold**

Supplementary Table 7: Optimal classification threshold for the best classifier of each feature set in each language determined through ROC analysis

| | **Feature set optimal classification threshold** | | | | |
|---|---|---|---|---|---|
| **Language** | Image-Text Congruence | Speech-timing | Word Params | Linguistic | Ensemble |
| NDL | 0.79 | 0.10 | 0.62 | 0.95 | 0.91 |
| DL | 1 | 0.71 | 0.91 | 0.91 | 0.73 |
| Spanish | 0.54 | 0.74 | 0.59 | 0.91 | 0.57 |
| Catalan | 0.66 | 0.77 | 0.90 | 0.73 | 0.79 |

Supplementary Table 8: classification performance for each classification algorithm in the non-dominant language. The classification performance of each feature set's best-performing classification algorithm is bolded. * indicates tie broken by AUC score.

| | **Feature set F1 macro** | | | | |
|---|---|---|---|---|---|
| **Classification Algorithm** | Image-Text Congruence | Speech-timing | Word Params | Linguistic | Ensemble |
| Decision Tree | **86** | **80** | 69 | **73** | **85** |
| Gradient Boosting | **86*** | 70 | 74 | 69 | **86** |
| Support Vector Machine | 81 | 77 | **93** | 50 | 81 |
| Shallow Neural Net | 81 | 72 | 61 | 23 | 74 |
| MAX | **86** | **80** | **85** | **73** | **86** |

Supplementary Table 9: Mean, standard deviation (SD), and statistical comparisons of word-level parameters across language dominance and subtype. Significant p-values are bolded.

| | nfv | | | lv | | | NDL | DL |
|---|---|---|---|---|---|---|---|---|
| **Feature** | **NDL (Mean, SD)** | **DL (Mean, SD)** | **p** | **NDL (Mean, SD)** | **DL (Mean , SD)** | **p** | **nfvPPA vs lvPPA p-value** | |
| Frequency per million* | 9010, 1650 | 9440, 2540 | 1 | 9400, 1400 | 8300, 1400 | **.014** | .76 | .14 |
| Verb frequency per million* | 1530, 1710 | 1670, 1830 | 1 | 870, 1300 | 2400, 1400 | **.001** | .76 | .18 |
| Noun frequency per million* | 191, 78.2 | 220, 120 | 1 | 200, 70 | 290, 90 | **.001** | .84 | .09 |
| Proportion high-frequency verbs | 0.357, 0.123 | 0.379, 0.194 | 1 | 0.423, 0.156 | 0.5, 0.1 | .06 | .62 | .07 |
| Word length by syllables* | 1.67, 0.118 | 1.67, 0.150 | 1 | 1.65, 0.107 | 1.57, 0.086 | **.01** | .76 | .06 |
| Concreteness* | 3.12, 0.079 | 3.09, 0.10 | 1 | 3.0, 0.06 | 2.97, 0.060 | 0.06 | **<.001** | **<.001** |
| Age of acquisition* | 2.23, 1.20 | 2.2, 1.1 | 1 | 1.61, 0.91 | 2.68, 1.02 | **.005** | .62 | .34 |
| Word length by phonemes* | 3.47, 0.364 | 3.5, 0.5 | 1 | 3.49, 0.33 | 3.25, 0.22 | **0.01** | .84 | .09 |

*averaged across each word

Supplementary Table 10: Mean, standard deviation (SD), and statistical comparisons of speech-timing measures across language dominance and subtype. Significant p-values are bolded.

| | **nfv** | | | **lv** | | | **NDL** | **DL** |
|---|---|---|---|---|---|---|---|---|
| **Feature** | **NDL (Mean, SD)** | **DL (Mean, SD)** | **p** | **NDL (Mean, SD)** | **DL (Mean, SD)** | **p** | **nfvPPA vs lvPPA p-value** | |
| Mean pause duration | 1.2, 0.55 | 0.90, 0.40 | .46 | 1.0, 0.36 | 1.0, 0.42 | .91 | .78 | .51 |
| Variability of pause duration | 0.90, 0.88 | 0.40, 0.40 | .46 | 0.69,0.71 | 0.70, 0.71 | .91 | .66 | .41 |
| Speech rate | 2.0, 0.36 | 2.3, 0.53 | .46 | 2.62, 0.64 | 2.75, 0.70 | .88 | **.02** | .14 |
| Average syllable duration | 0.30, 0.06 | 0.29, 0.05 | .94 | 0.24, 0.03 | 0.24, 0.03 | .88 | **.002** | **.009** |
| Articulation rate | 3.49, 0.63 | 3.5, 0.60 | .94 | 4.2, 0.43 | 4.3, 0.48 | .88 | **.002** | **.004** |
| Speech-to-pause ratio | 1.65, 0.97 | 14, 34 | .46 | 2.8, 4.8 | 11, 44.1 | .88 | .65 | .84 |
| # pauses | 42.5, 22.2 | 33, 25 | .46 | 51, 27 | 49, 37 | .88 | .65 | .29 |
| # words/min | 55, 29 | 68, 31 | .46 | 73, 26 | 93, 39 | .25 | **.03** | .14 |
| # propositions/min | 20, 13 | 26, 13 | .46 | 32, 14 | 43, 19 | .25 | **.02** | **.04** |

Supplementary Table 11: Mean, standard deviation (SD), and statistical comparisons of linguistic features across language dominance and subtype. Significant p-values are bolded. Marginally significant (0.05<p<0.1) p-values are italicized.

| | **nfv** | | | **lv** | | | **NDL** | **DL** |
|---|---|---|---|---|---|---|---|---|
| **Feature** | **NDL (Mean, SD)** | **DL (Mean, SD)** | **p** | **NDL (Mean, SD)** | **DL (Mean, SD)** | **p** | **nfvPPA vs lvPPA p-value** | |
| # words | 90, 55 | 104, 90 | .87 | 154, 79 | 183, 108 | .58 | **.03** | **.009** |
| Type token ratio | 0.57, 0.07 | 0.58, 0.13 | .87 | 0.54, 0.08 | 0.51, 0.08 | .39 | .61 | *.08* |
| Noun to verb ratio | 2.0, 0.74 | 1.5, 0.63 | .56 | 1.4, 0.51 | 1.1, 0.40 | .27 | *.05* | *.07* |
| Propositional Density | 0.37, 0.06 | 0.39, 0.09 | .74 | 0.43, 0.06 | 0.46, 0.05 | .27 | **.03** | **.009** |
| Open-closed words ratio | 0.74, 0.14 | 0.83, 0.17 | .56 | 0.73, 0.11 | 0.78, 0.11 | .27 | .84 | .42 |
| Average sentence length | 8.94, 4.4 | 7.4, 2.6 | .56 | 9.3, 2.3 | 9.9, 2.2 | .58 | .52 | **.009** |
| Subordination main verb implied | 0.38, 0.30 | 0.26, 0.29 | .56 | 0.59, 0.28 | 0.69, 0.27 | .54 | .11 | **.009** |
| Proportion fillers | 0.07, 0.05 | 0.05, 0.06 | .56 | 0.07, 0.05 | 0.05, 0.03 | .58 | .86 | .43 |
| Retracing | 0.03, 0.03 | 0.05, 0.04 | .56 | 0.07, 0.05 | 0.07, 0.05 | .74 | *.06* | .19 |
| Whole-word repetition | 0.12, 0.17 | 0.09, 0.09 | .87 | 0.09, 0.07 | 0.08, 0.06 | .58 | .84 | .92 |
| Part-word repetition | 0.04, 0.04 | 0.03, 0.03 | .94 | 0.03, 0.02 | 0.03, 0.02 | .74 | .84 | .43 |

Supplementary Table 12: Classification performance for each classification algorithm in the dominant language. The classification performance of each feature set's best-performing classification algorithm is bolded. * indicates tie broken by AUC score.

| | **Feature set F1 macro** | | | | |
|---|---|---|---|---|---|
| **Classification Algorithm** | Image-Text Congruence | Speech-timing | Word Params | Linguistic | Ensemble |
| Decision Tree | 66 | 53 | **87** | 79 | 80 |
| Gradient Boosting | 69 | 67 | 82 | **87** | **92** |
| Support Vector Machine | 69 | 70 | 56 | 72 | 84 |
| Shallow Neural Net | **82** | **70*** | 56 | 77 | 84 |
| MAX | **82** | **70** | **87** | **87** | **92** |

Supplementary Table 13: Mean, SD, statistical comparisons, permutation analysis of each vision-language encoder's image-text congruence score across language dominance and subtype. Significant p-values are bolded. Marginally significant (0.05<p<0.1) p-values are italicized.

| | **nfv** | | | **lv** | | | **NDL** | **DL** |
|---|---|---|---|---|---|---|---|---|
| **Vision-language encoder** | **NDL (Mean, SD)** | **DL (Mean, SD)** | **p** | **NDL (Mean, SD)** | **DL (Mean, SD)** | **p** | **nfvPPA vs lvPPA p-value, permutation importance (%)** | |
| ViT-B-16-plus-240 paired with M-CLIP/XLM-Roberta-Large-Vit-B-16Plus | 0.32, 0.08 | 0.31, 0.06 | .80 | 0.25, 0.05 | 0.26, 0.05 | .83 | **<.001**, 41 | **0.03**, 12 |
| ViT-B/32 paired with M-CLIP/XLM-Roberta-Large-Vit-B-32 | 0.28, 0.03 | 0.28, 0.03 | .80 | 0.25, 0.02 | 0.25, 0.02 | .83 | **.004**, 1 | *0.05*, 6 |
| ViT-L/14 paired with M-CLIP/LABSE-Vit-L-14 | 0.20, 0.02 | 0.19, 0.02 | .80 | 0.21, 0.02 | 0.20, 0.02 | *.06* | **.04**, 0 | 0.69, 1 |
| ViT-L/14 paired with M-CLIP/XLM-Roberta-Large-Vit-L-14 | 0.29, 0.05 | 0.28, 0.05 | .80 | 0.25, 0.03 | 0.24, 0.05 | .83 | **.003**, 0 | *0.05*, 1 |

Supplementary Table 14: Mean, standard deviation (SD), and statistical comparisons of part-of-speech tags relevant to propositional density across language dominance and subtype. Significant p-values are bolded.

| | **nfv** | | | **lv** | | | **NDL** | **DL** |
|---|---|---|---|---|---|---|---|---|
| **Feature** | **NDL (Mean, SD)** | **DL (Mean, SD)** | **p** | **NDL (Mean, SD)** | **DL (Mean, SD)** | **p** | **nfvPPA vs lvPPA p-value** | |
| Verb | 0.14, 0.04 | 0.18, 0.04 | .22 | 0.15, 0.03 | 0.17, 0.03 | .15 | .59 | .51 |
| Adjective | 0.02, 0.02 | 0.02, 0.02 | .85 | 0.02, 0.02 | 0.02, 0.01 | .89 | .59 | .51 |
| Adverb | 0.04, 0.03 | 0.03, 0.05 | .85 | 0.09, 0.04 | 0.10, 0.03 | .81 | **.006** | **.002** |
| Adposition | 0.08, 0.05 | 0.08, 0.04 | .85 | 0.09, 0.03 | 0.09, 0.02 | .89 | .66 | .48 |
| Subordinating conjunction | 0.02, 0.02 | 0.01, 0.01 | .85 | 0.03, 0.02 | 0.03, 0.01 | .88 | .59 | **.02** |
| Coordinating conjunction | 0.07, 0.04 | 0.08, 0.04 | .85 | 0.06, 0.02 | 0.06, 0.02 | .89 | .59 | .24 |

**Comparing classification performance between Spanish and Catalan**

We evaluated the classification performance of our approach between samples in Spanish (a language where NLP tools are well developed; high-resource language, and Catalan (a language where NLP tools are underdeveloped; low-resource language[67]). If we could derive features in both languages to a similar level of robustness, then we predict that our classifier trained on Spanish samples would perform similarly to our classifier trained on Catalan samples. This was indeed the case as the classification performance between either language was not significantly different for any of the feature sets or the ensemble (p=1 for each feature set's classifier and the ensemble classifier, Supplementary Table 15). This is evidence that we robustly derived features in both languages, and more generally that this approach can also differentiate nfvPPA from lvPPA in low-resource (Catalan) languages.

Supplementary Table 15: Comparing classification performance between Spanish and Catalan for each feature set and the ensemble classifier

| Feature set | Spanish F1 macro | Catalan F1 macro | McNemar p |
|---|---|---|---|
| Image-text congruence | 89 | 85 | 1 |
| Speech timing | 75 | 76 | 1 |
| Word params | 85 | 87 | 1 |
| Linguistic | 86 | 74 | 0.45 |
| Ensemble | 86 | 87 | 1 |

Additionally, the classification performance of each classification algorithm is included for completeness (Supplementary Tables 16 and 17).

Supplementary Table 16: Percent F1 macro classification performance for each classification algorithm in the Spanish language. The classification performance of each feature set's best-performing classification algorithm is bolded.

| | **Feature set F1 macro (%)** | | | | |
|---|---|---|---|---|---|
| **Classification Algorithm** | Image-Text Congruence | Speech-timing | Word Params | Linguistic | Ensemble |
| Decision Tree | 85 | 73 | 61 | **86** | 77 |
| Gradient Boosting | **89** | 74 | 72 | 74 | 79 |
| Support Vector Machine | 87 | **75** | **85** | 61 | **86** |
| Shallow Neural Net | 84 | 64 | 23 | 74 | 84 |
| MAX | **89** | **75** | **85** | **86** | **86** |

Supplementary Table 17: Percent F1 macro classification performance for each classification algorithm in the Catalan language. The classification performance of each feature set's best-performing classification algorithm is bolded.

| | **Feature set F1 macro (%)** | | | | |
|---|---|---|---|---|---|
| **Classification Algorithm** | Image-Text Congruence | Speech-timing | Word Params | Linguistic | Ensemble |
| Decision Tree | 82 | 64 | **87** | 70 | 74 |
| Gradient Boosting | **85** | 63 | 82 | **74** | **87** |
| Support Vector Machine | 82 | **76** | 56 | 67 | 83 |
| Shallow Neural Net | 67 | 53 | 56 | 73 | 85 |
| MAX | **85** | **75** | **86** | **74** | **87** |
